# Hospital Price Transparency Data Reveal Up to 8-Fold Geographic Variation in Commercial Rates for IR Procedures

**DOI:** 10.64898/2026.05.09.26352821

**Authors:** Pouyan Golshani, Mary Joseph

## Abstract

**Objective:** To characterize the magnitude and geographic distribution of commercially negotiated hospital facility rates for fourteen common interventional radiology (IR) procedures using publicly posted Hospital Price Transparency Machine-Readable Files (MRFs), and to describe the relationships between state-level commercial pricing, population rurality, and within-system rate uniformity.

**Methods:** In this cross-sectional observational analysis, we examined hospital-weighted commercial rate observations from U.S. hospital MRFs for fourteen IR procedures spanning image-guided drainage, embolization, peripheral vascular intervention, dialysis access maintenance, and percutaneous spine. The unit of analysis was one observation per distinct negotiated rate per state-CPT cell, deduplicating multi-facility same-system reporting in which two or more hospitals posted identical rate, range, and payer-count tuples. Outliers were excluded using transparent absolute and CMS-relative bounds. State-level statistics were computed where ≥5 distinct hospital-system observations were reported. Commercial rates were compared to CY 2026 CMS Outpatient Prospective Payment System (OPPS) facility payments. Relationships between state-level commercial rate and 2020 U.S. Census percent-rural population were assessed by Spearman rank correlation.

**Results:** Across 14 procedures, state-level commercial median rates varied 3.7-to 8.3-fold between the highest- and lowest-priced states. The largest spreads were observed for fem-pop angioplasty (CPT 37224, 8.3-fold), fem-pop atherectomy (37225, 8.1-fold), and iliac stenting (37221, 7.1-fold). National median commercial rates ranged from 1.34× (PAE/GAE) to 3.60× (paracentesis) the corresponding CMS OPPS facility payment. Across all 14 procedures, the relationship between state percent-rural and median commercial rate was negative (mean Spearman ρ = −0.46, range −0.33 to −0.80; 14 of 14 codes negative), with the most-rural quartile of states showing a median commercial rate 42% below the most-urban quartile. Deduplication identified 660 multi-facility groups in which a single negotiated rate was applied across two or more affiliated hospitals within a state.

**Discussion:** Substantial state-level variation in commercially negotiated facility rates exists for common IR procedures, with consistently lower rates in more rural states. Within-system rate uniformity is a frequent feature: many regional health systems post identical commercial rates across multiple owned facilities. The findings are consistent with prior literature linking commercial pricing to market structure and support continued investment in price transparency as a precondition for informed decision-making.

## Introduction

In 2021, the Centers for Medicare & Medicaid Services (CMS) Hospital Price Transparency Final Rule (45 CFR Part 180) required all U.S. hospitals to publish a machine-readable file listing standard charges for items and services across all payers and plans. The intent was to make commercial reimbursement, historically opaque under non-disclosure provisions, accessible to patients, employers, payers, researchers, and clinicians. Subsequent analyses of these data have demonstrated wide variation in posted commercial rates across hospitals and metropolitan areas for common surgical and medical procedures, alongside ongoing concerns about inconsistent posting compliance. Comprehensive Machine-Readable File-based analyses focused specifically on interventional radiology procedures have to date been limited.

Interventional radiology occupies a distinctive position in modern hospital-based care: a single specialty performs procedures spanning image-guided drainage, embolization for benign and malignant disease, peripheral vascular revascularization, dialysis access maintenance, percutaneous spine intervention, and central venous access. Many of these procedures can be performed in inpatient, hospital outpatient, or ambulatory surgery settings. Patients, referring physicians, and operators increasingly need transparent rate data to inform care setting choices and value-based contracting decisions.

In this study we use Machine-Readable File data, deduplicated and hospital-weighted at the system level, to describe the magnitude and direction of state-level variation in commercial facility rates for fourteen common IR procedures. We compare commercial rates to CY 2026 CMS Outpatient Prospective Payment System payments as a stable reference, examine the relationship between state-level commercial pricing and population rurality, and characterize the prevalence of within-system rate uniformity. Our purpose is descriptive: to make the variation visible and characterize its patterns, leaving inferences about cost structure or policy implication to readers.

## Methods

### Study design

Cross-sectional observational analysis of publicly posted U.S. hospital Machine-Readable File commercial rate data, with comparison to CMS facility payment rates and to U.S. Census-derived state population characteristics. Reported in accordance with the STROBE statement. The study used only de-identified, publicly available data and was exempt from human subjects review.

### Data sources

Commercial rate data were drawn from CenterIQ (https://gighz.com/centeriq/), a curated database of U.S. hospital Machine-Readable Files maintained by GigHz (https://gighz.com) that standardizes negotiated commercial facility rates at the payer-arrangement level and aggregates them to per-hospital median rates per Current Procedural Terminology (CPT®) code. CMS Outpatient Prospective Payment System (OPPS) payment rates were obtained from the CY 2026 January Web Addendum B (released December 29, 2025). For procedures paid under Comprehensive APC (C-APC) methodology, where individual code line payment rates are not separately listed, the mapped APC tier from the CY 2026 OPPS Final Rule was used as the comparator. Population rurality data were obtained from the 2020 U.S. Decennial Census (percent of state population residing in rural areas).

### Procedure selection

Fourteen IR procedures were selected to span the major categories of contemporary practice: image-guided drainage and biopsy guidance, percutaneous biliary drainage, percutaneous spine, embolization, peripheral vascular diagnostic and interventional procedures, and dialysis access intervention. Two image-guidance/add-on codes (CPT 10006, 77002) were excluded from the headline analysis and presented in a separate sensitivity panel because their extreme observed variation in MRFs most likely reflects posting heterogeneity rather than true negotiated rate variation.

### Unit of observation and within-system deduplication

To prevent inflation of variation estimates from same-system multi-facility reporting, we deduplicated within each state-CPT cell at the negotiated-rate level: rows sharing identical rate_median, rate_min, rate_max, and payer_count tuples were collapsed to a single observation. This treats each distinct negotiated commercial rate as one observation regardless of how many affiliated facilities (e.g., multiple campuses of a regional health system, freestanding emergency departments, or affiliated urgent-care entities) post that rate. Entities matching name patterns indicating freestanding emergency departments or ambulance services, which do not perform the analyzed procedures but inherit parent-system rates, were excluded prior to deduplication. State-level statistics were computed where ≥5 distinct (state-CPT) observations remained after deduplication and filtering.

### Outlier filtering

Hospital observations were filtered using: (1) absolute hard bounds excluding rates < $25 or > $250,000; (2) where a CMS OPPS payment rate was available, code-relative bounds excluding rates outside [0.25 × CMS_OPPS, 6.0 × CMS_OPPS], consistent with prior MRF research methodology; (3) state-level small-cell suppression for cells with <5 distinct observations; and (4) state-level exclusion of Hawaii due to small reporting hospital count and atypical hospital structure that produced rates outside expected ranges across nearly all study codes. Sensitivity analyses without code-relative outlier filtering yielded substantively unchanged headline findings.

### Statistical analysis

State-level rates were summarized by median, 25th and 75th percentiles. Geographic variation was characterized by the highest-state-to-lowest-state ratio of state median rates. Commercial-to-CMS ratios were computed as state median divided by CY 2026 OPPS APC payment. The relationship between state median commercial rate and percent-rural population was assessed by Spearman rank correlation; rural-versus-urban quartile comparisons compared the median state-median rate of the most-rural quartile to that of the most-urban quartile of reporting states.

### Important methodological clarification: facility versus professional fee

All rates analyzed reflect commercial facility (technical) payments to hospitals. Physician professional fees, billed under separate CPT modifiers and reimbursed under the CMS Physician Fee Schedule, are negotiated separately with commercial payers and historically show much narrower geographic variation than facility rates. Findings should not be interpreted as reflecting variation in physician earnings.

## Results

### Sample characteristics

After application of outlier and small-cell filters, exclusion of Hawaii, removal of non-procedural facility entities (n = 294 entities, predominantly freestanding emergency departments and ambulance services inheriting parent-system rates), and within-state deduplication of identical-rate multi-facility groups (n = 660 groups collapsed across 14 codes), the analytic sample contained 3,856 distinct facility-rate observations across the 14 headline procedures. The number of states meeting the 5-observation reporting threshold ranged from 16 (CPT 49083 paracentesis) to 31 (CPT 36906 dialysis circuit revision).

### Geographic variation

Across the 14 headline procedures, the highest-state-to-lowest-state ratio of state-median commercial rates ranged from 3.7× (CPT 36561 tunneled port) to 8.3× (CPT 37224 fem-pop angioplasty) (Table 1). Median state commercial rates ranged from $3,334 (paracentesis) to $39,952 (fem-pop stent + atherectomy). The states most frequently anchoring the high end across multiple procedures were Pennsylvania, Indiana, New Jersey, Florida, and California; states most frequently anchoring the low end were Maine, North Dakota, Utah, New Hampshire, and South Carolina.

**Figure 1.**
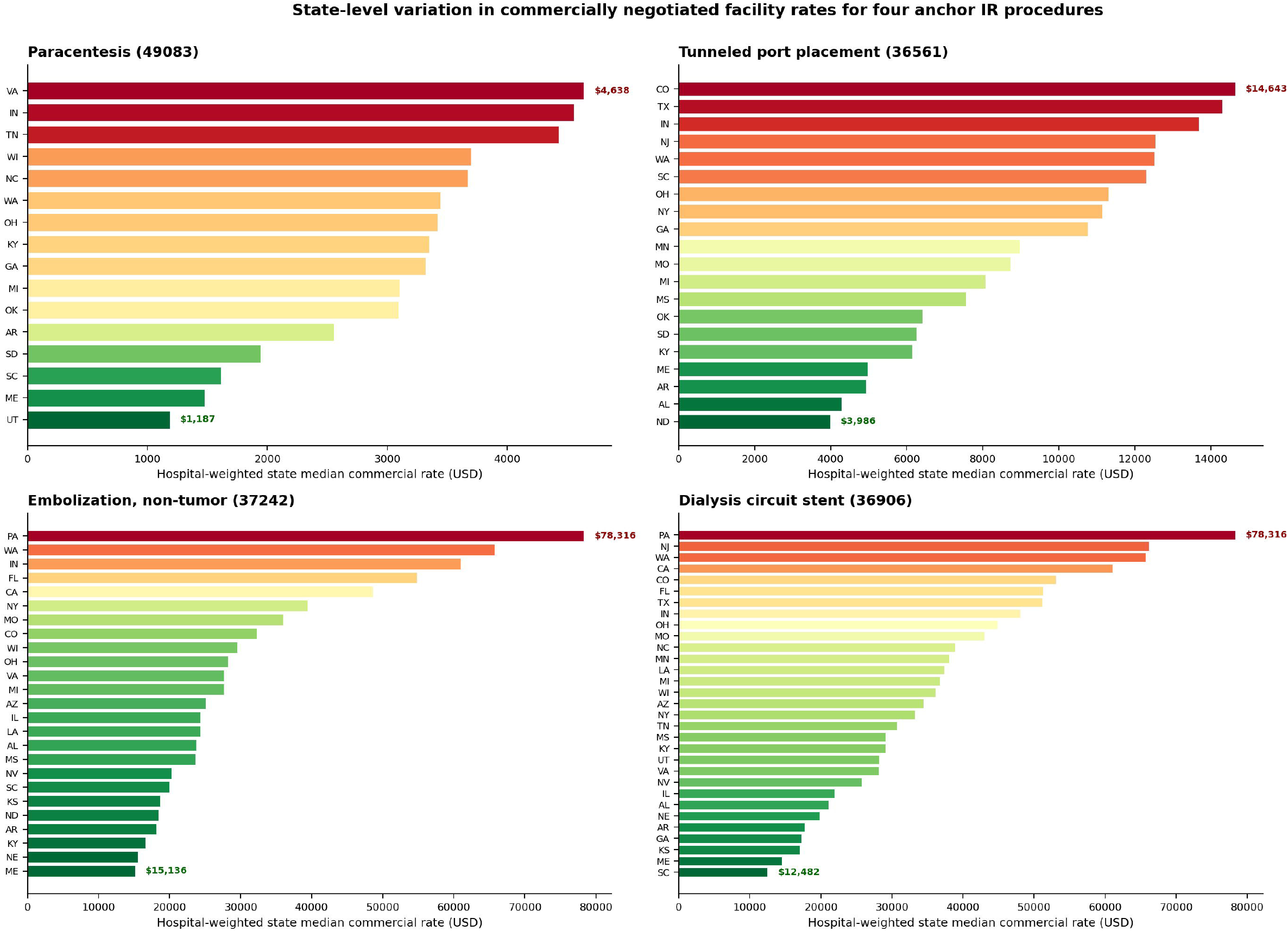
Hospital-system-deduplicated state-level commercial rate distribution for four anchor IR procedures: paracentesis with imaging (CPT 49083), tunneled port placement (36561), embolization for non-tumor/non-hemorrhage indication (37242), and dialysis circuit revision with stent (36906).

**Table 1.** State-level commercial rate distributions for 14 IR procedures (system-deduplicated, hospital-weighted)

| CPT | Procedure | States | P25 | Median | P75 | Lowest state | Highest state |
| --- | --- | --- | --- | --- | --- | --- | --- |
| 37224 | Fem-pop revascularization, angioplasty | 23 | \$12,065 | **\$17,167** | \$24,169 | ME:\$4,758 | NY:\$39,385 |
| 37225 | Fem-pop revascularization, atherectomy | 25 | \$21,844 | **\$27,299** | \$39,732 | ME:\$9,667 | PA:\$78,316 |
| 37221 | Iliac revascularization, with stent | 27 | \$20,060 | **\$29,808** | \$43,533 | NH:\$11,202 | IN:\$80,016 |
| 47533 | Percutaneous biliary drain, internal-external | 28 | \$5,895 | **\$8,398** | \$11,145 | ME:\$2,314 | PA:\$15,452 |
| 36906 | Endovascular dialysis circuit revision, with stent | 31 | \$21,934 | **\$34,457** | \$48,045 | SC:\$12,482 | PA:\$78,316 |
| 37231 | Tibial-peroneal revascularization, stent + atherectomy | 24 | \$24,457 | **\$39,070** | \$49,847 | ME:\$13,250 | PA:\$84,132 |
| 37226 | Fem-pop revascularization, with stent | 24 | \$21,571 | **\$27,868** | \$39,385 | ME:\$12,167 | IN:\$72,586 |
| 37227 | Fem-pop revascularization, stent + atherectomy | 24 | \$29,474 | **\$39,952** | \$53,226 | ME:\$13,404 | PA:\$80,571 |
| 75710 | Lower-extremity angiography, unilateral | 18 | \$5,452 | **\$8,601** | \$10,535 | ME:\$2,809 | FL:\$15,541 |
| 37242 | Embolization, non-tumor / non-hemorrhage (PAE, GAE) | 25 | \$19,984 | **\$25,080** | \$35,994 | ME:\$15,136 | PA:\$78,316 |
| 49083 | Abdominal paracentesis with imaging guidance | 16 | \$2,555 | **\$3,334** | \$3,696 | UT:\$1,187 | VA:\$4,638 |
| 22513 | Vertebral augmentation (kyphoplasty), thoracic | 25 | \$12,262 | **\$14,524** | \$19,145 | NV:\$9,650 | CA:\$37,984 |
| 37243 | Embolization for tumor / organ ischemia (UFE, TARE) | 27 | \$15,785 | **\$19,736** | \$25,201 | GA:\$13,300 | FL:\$51,008 |
| 36561 | Tunneled central venous catheter with subcutaneous port | 20 | \$6,251 | **\$8,850** | \$12,518 | ND:\$3,986 | CO:\$14,643 |

Note: P25, median, and P75 are computed across state-level medians (distribution of states), not across hospitals.

### Within-system rate uniformity

Deduplication revealed widespread within-system rate uniformity. The procedure flagged 660 multi-facility groups across the 14 codes, in which two or more affiliated hospitals within a state posted exactly identical commercial rate, range, and payer-count tuples. Examples included AdventHealth Orlando (8 facilities posting identical rates for fem-pop revascularization with stent + atherectomy), Honor Health Arizona (7 facilities for percutaneous biliary drain), MultiCare Washington (5 facilities for iliac stenting), Capital Health New Jersey (5 facilities for several codes), and the Lutheran Health Network of Fort Wayne, Indiana (multiple codes). This pattern—where a single negotiated commercial rate is applied across an entire health system’s facility list, regardless of facility size, type, or referral status—was observed in every state with ≥3 reporting facilities for at least one code.

### Commercial relative to Medicare

For the 8 of 14 procedures with a published CY 2026 OPPS APC payment rate, commercial state-median rates ranged from 1.34× (CPT 37242 non-tumor embolization) to 3.60× (CPT 49083 paracentesis) the OPPS facility payment (Table 2). Paracentesis (3.60×), tunneled port (2.74×), lower-extremity angiography (2.67×), and biliary drain (2.30×) showed the largest commercial-to-Medicare premiums.

**Figure 2.**
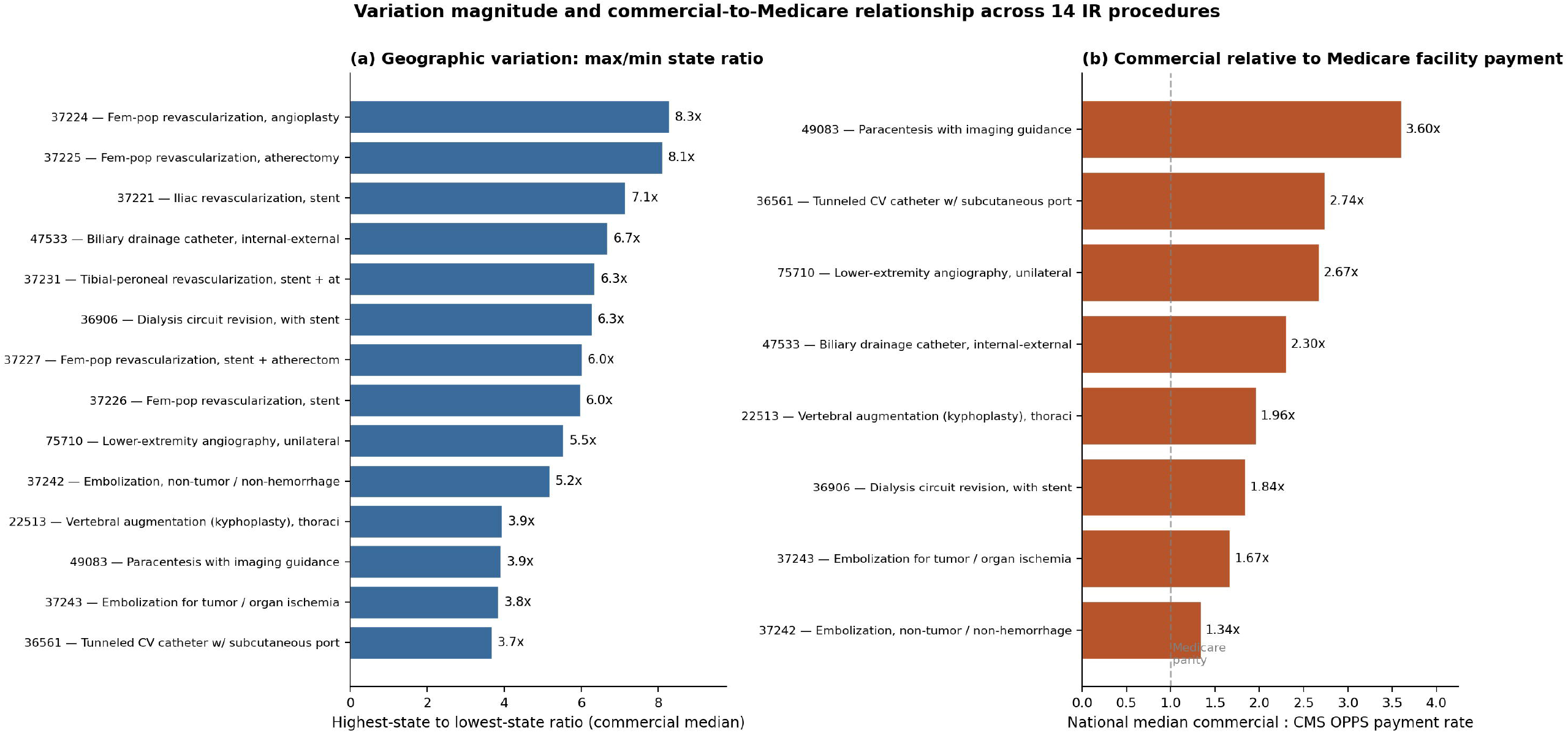
(a) Highest-state to lowest-state ratio of state median commercial rates by procedure. (b) National median commercial rate as a multiple of CY 2026 CMS OPPS facility payment, where available.

**Table 2.**
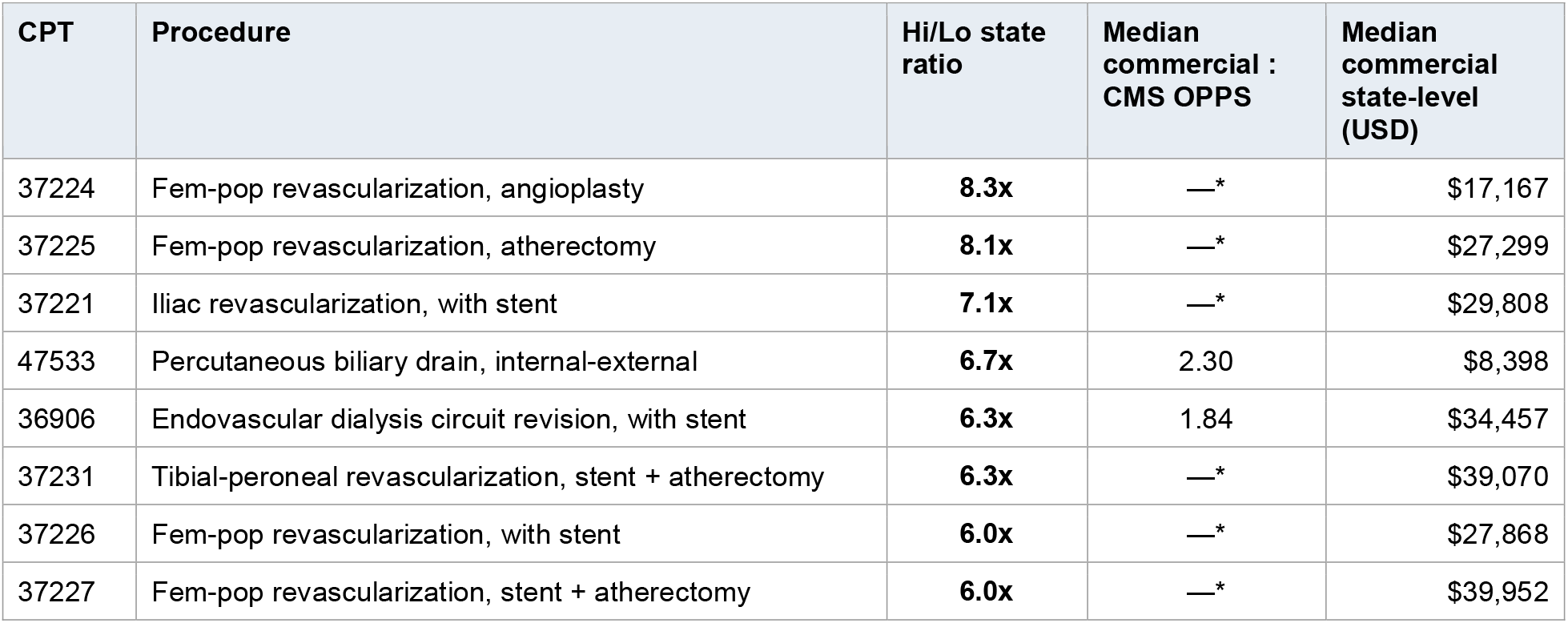
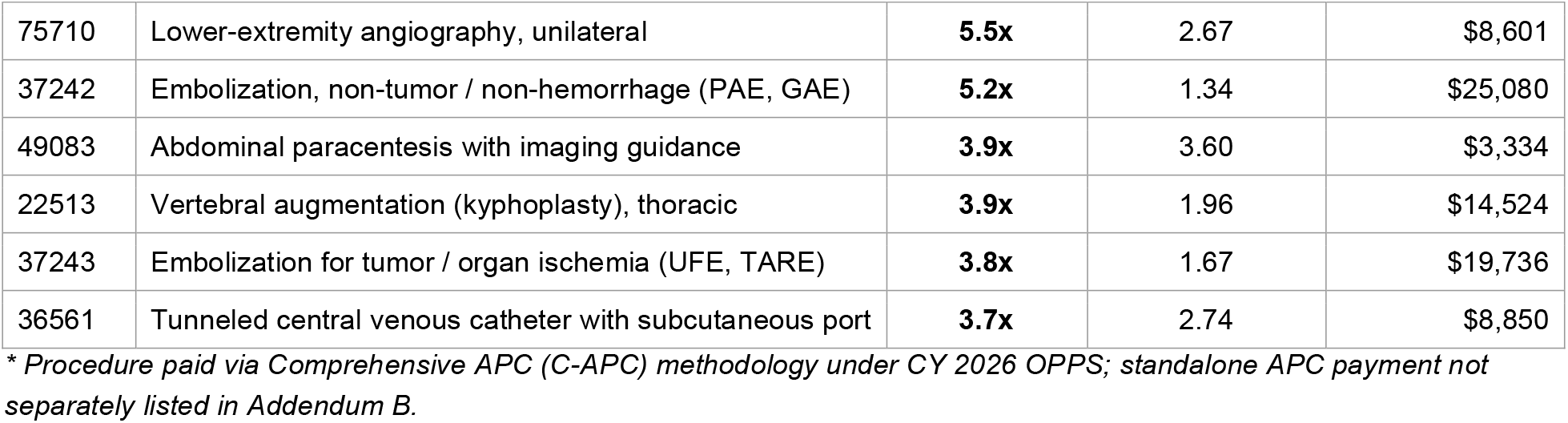
Commercial median rates relative to CY 2026 CMS OPPS facility payment.

### State percent-rural and commercial rate

Across all 14 headline procedures, the relationship between state percent-rural population and state-median commercial rate was negative (mean Spearman ρ = −0.46, range −0.33 to −0.80; 14 of 14 codes negative; Table 3). The most-rural quartile of states had a median commercial rate 42% below the most-urban quartile, averaged across the 14 codes.

**Figure 3.**
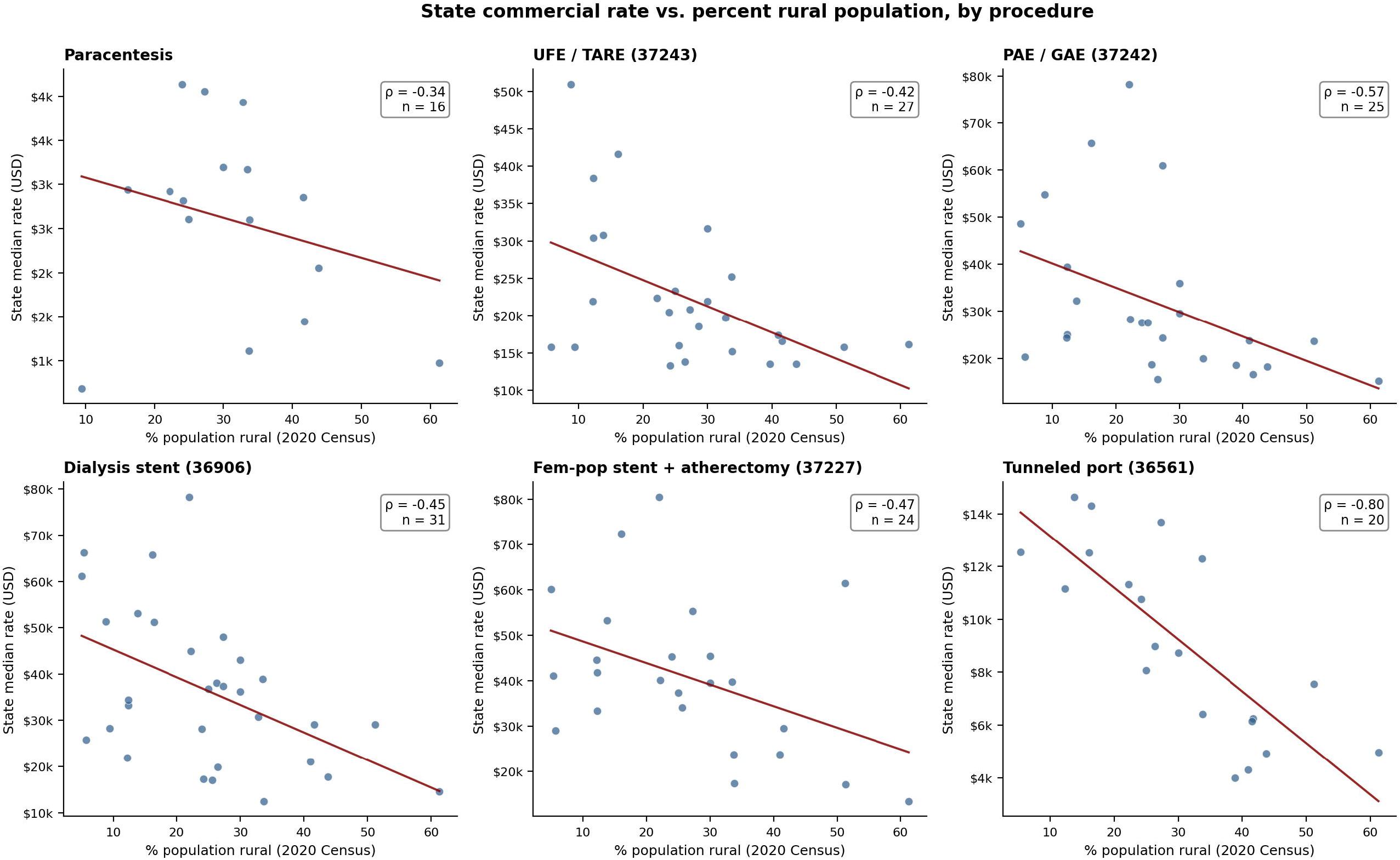
Relationship between state population percent-rural (2020 U.S. Census) and state-median commercial rate for six representative IR procedures, with linear fit and Spearman correlation. The negative relationship is consistent across procedures.

**Table 3.** State commercial rate vs. percent-rural population.

| CPT | Procedure | States | Spearman $\rho$ | Urban-Q median (USD) | Rural-Q median (USD) | Rural – Urban (%) |
| --- | --- | --- | --- | --- | --- | --- |
| 36561 | Tunneled central venous catheter with subcutaneous port | 20 | **-0.798** | \$12,550 | \$6,142 | -51.1% |
| 37242 | Embolization, non-tumor / non-hemorrhage (PAE, GAE) | 25 | **-0.570** | \$32,259 | \$18,492 | -42.7% |
| 37227 | Fem-pop revascularization, stent + atherectomy | 24 | **-0.473** | \$41,460 | \$20,540 | -50.5% |
| 37224 | Fem-pop revascularization, angioplasty | 23 | **-0.470** | \$29,049 | \$10,761 | -63.0% |
| 37225 | Fem-pop revascularization, atherectomy | 25 | **-0.452** | \$34,641 | \$20,332 | -41.3% |
| 36906 | Endovascular dialysis circuit revision, with stent | 31 | **-0.452** | \$33,253 | \$25,079 | -24.6% |
| 75710 | Lower-extremity angiography, unilateral | 18 | **-0.451** | \$9,434 | \$5,289 | -43.9% |
| 37231 | Tibial-peroneal revascularization, stent + atherectomy | 24 | **-0.451** | \$37,182 | \$24,075 | -35.3% |
| 47533 | Percutaneous biliary drain, internal-external | 28 | **-0.439** | \$9,450 | \$5,757 | -39.1% |
| 37243 | Embolization for tumor / organ ischemia (UFE, TARE) | 27 | **-0.422** | \$26,180 | \$15,785 | -39.7% |
| 37226 | Fem-pop revascularization, with stent | 24 | **-0.418** | \$34,178 | \$19,412 | -43.2% |
| 22513 | Vertebral augmentation (kyphoplasty), thoracic | 25 | **-0.372** | \$19,177 | \$12,262 | -36.1% |
| 49083 | Abdominal paracentesis with imaging guidance | 16 | **-0.344** | \$3,431 | \$2,250 | -34.4% |
| 37221 | Iliac revascularization, with stent | 27 | **-0.333** | \$30,915 | \$18,451 | -40.3% |

## Discussion

This analysis, based on deduplicated, hospital-system-weighted Machine-Readable File data for 14 common IR procedures, demonstrates substantial state-level variation in commercially negotiated facility rates—ranging from approximately 4-to 8-fold between the highest and lowest state—and a consistent inverse relationship between state percent-rural population and commercial rate magnitude. We additionally observe that commercial rates frequently exhibit within-system uniformity, with many regional health systems posting identical negotiated rates across all owned facilities. These findings are descriptive; they do not by themselves indicate that prices in any state, hospital, or system are too high or too low. They do make visible patterns previously documented in the broader hospital-pricing literature, with direct relevance to IR practice and patient care.

Several features of hospital cost structure and market environment likely contribute to the patterns observed. Larger metropolitan and academic centers serve patient populations with higher acuity and disproportionate uncompensated-care burden; these costs may be cross-subsidized through commercial payment, particularly for hospitals serving as regional referral centers. Capital expenditures, labor costs, and physician compensation also vary by metropolitan area. Negotiated commercial rates, once established, often persist for years and reflect the bargaining circumstances of one historical point in time rather than current cost. None of these factors makes geographic variation inappropriate; they make it complex.

The inverse relationship between state percent-rural and commercial rate is consistent with the market-structure literature on hospital pricing, including work by Cooper and colleagues showing that commercial prices vary more strongly with hospital market concentration than with cost.

Concentrated regional health networks—where one or a small number of systems dominate the available choice set for commercial enrollees—have negotiating leverage that less consolidated markets lack. Our within-state observations, including the Indiana finding that high-rate clustering tracks specific regional networks, are consistent with this pattern.

The within-system rate uniformity finding has practical implications for patients and clinicians. When a single negotiated commercial rate applies across all facilities owned by a regional system—from large academic referral centers to small community hospitals to acquired freestanding emergency departments—patients selecting among in-system facilities encounter no rate variation regardless of facility type. As regional acquisition continues, the geographic distribution of commercial rates will increasingly reflect post-acquisition system pricing rather than legacy facility-level pricing. Both consolidated and independent operators have a vested interest in healthcare delivery remaining sustainable; the broader question is whether all participants in the system can see the prices that apply to the care they deliver, refer for, or pay for.

It is also notable that across all 8 procedures with a published CY 2026 OPPS payment rate, the median state commercial rate exceeded the corresponding Medicare facility payment, in some cases by more than 3-fold. Medicare, as the largest single payer in the United States, effectively functions as the floor of facility reimbursement for most hospital-based procedures. Commercial rates negotiated above this floor reflect the bargaining outcomes of individual hospital-payer pairs and the market structures in which those negotiations occur.

It is important to emphasize what this study does not measure. Commercial facility rates posted in MRFs are negotiated payments, not charges and not what individual patients ultimately pay. They reflect facility (technical) reimbursement only; physician professional fees—what the interventional radiologist or other physicians actually receive—are negotiated separately and historically show far less geographic variation. Conflation of facility rates with physician earnings is a common error in interpretation of MRF data and should be avoided.

### Limitations

MRF data quality is heterogeneous across hospitals and posting periods; despite filtering and deduplication, residual posting heterogeneity is likely. Our analysis is hospital-system-weighted, which is appropriate for descriptive variation analysis but does not capture patient-volume-weighted estimates. Six peripheral-vascular revascularization codes (CPT 37220 family) lack standalone CY 2026 OPPS payment rates because they are paid under Comprehensive APC methodology; commercial-to-Medicare ratio analysis for these codes is deferred to a subsequent paper using actual Medicare paid amounts. Our rurality measure is at the state level; future work should examine MSA-level patterns. This analysis is descriptive and does not attempt to attribute observed variation to any specific causal mechanism.

## Conclusion

Across 14 common interventional radiology procedures and 16 to 31 reporting states per procedure, commercially negotiated hospital facility rates show substantial state-level variation (3.7-to 8.3-fold between highest and lowest state), consistent inverse association with state-level rurality (mean Spearman ρ = −0.46), and frequent within-system rate uniformity in which a single negotiated rate is applied across an entire regional health system’s facility list. The patterns are consistent with prior literature on hospital market structure. We present these findings descriptively, with the goal of making the state of the commercial rate landscape visible to patients, employers, payers, clinicians, and policymakers. Continued investment in price transparency is the precondition for any of these participants to make informed decisions in the markets in which they operate.

## Data Availability

The underlying Hospital Price Transparency Machine-Readable Files are publicly available from individual U.S. hospitals as required by 45 CFR Part 180. The deduplicated state-level dataset analyzed in this manuscript is available from the corresponding author on reasonable request. The CenterIQ aggregated dataset is accessible at https://gighz.com/centeriq/.

https://www.cms.gov/medicare/medicare-fee-for-service-payment/hospitaloutpatientpps

